# An epidemiologic profile of COVID-19 patients in Vietnam

**DOI:** 10.1101/2020.04.10.20061226

**Authors:** Huy G. Nguyen, Tuan V. Nguyen

## Abstract

**Background and Aim:** There is a paucity of data on the COVID-19 pandemic in Vietnam. In this paper, we sought to provide an epidemiologic description of patients who were infected with SARS-Cov-2 in Vietnam.

**Methods:** Data were abstracted from the wikipedia’s COVID-19 information resource and Johns Hopkins University Dashboard. Demographic data and treatment status were obtained for each patient in each day. The coverage period was from 23/1/2020 to 10/4/2020. Descriptive analyses of incident cases were stratified by gender and age group. The estimation of the reproduction ratio was done with a bootstrap method using the R statistical environment.

**Results:** During the coverage period, Vietnam has recorded 257 cases of COVID-19. Approximately 54% of the cases were women. The median age of patients was 30 years (range: 3 months to 88 years), with 78% of patients aged 49 or younger. About 66% (n = 171) of patients were overseas tourists (20%) and Vietnamese students or workers returning from overseas (46%). Approximately 57% (n = 144) of patients have been recovered and discharged from hospitals. There have been no mortality. The reproduction ratio was estimated to range between 0.95 and 1.24.

**Conclusion:** These data indicate that a majority of COVID-19 patients in Vietnam was imported cases in overseas tourists and young students and workers who had returned from overseas.

## Introduction

The outbreak of the COVID-19 pandemic was initially reported in the city of Wuhan, Hubei Province, China. The outbreak was thought to be linked to a seafood market [1], with the causative pathogen being a novel betacoronavirus within the severe acute respiratory syndrome (SARS) coronavirus (CoV) family which is now known as SARS-CoV-2 [2]. The outbreak has since been spread to 185 countries and territories around the world. At the time of writing, the total number of confirmed cases was 1,611,981, with 96783 deaths being reported worldwide [3].

Vietnam, a neighbouring country of China, recorded the first case of COVID-19 on January 23, 2020. The case was a tourist from China, and the patient had been successfully treated in a tertiary hospital in Ho Chi Minh City. Since then, Vietnam has recorded more than more cases of COVID-19. However, there is little epidemiologic data on COVID-19 from Vietnam in the medical literature.

The primary aim of this short communication was therefore to provide a brief epidemiologic description of the COVID-19 situation in Vietnam. The report also provides an estimate of the reproduction ratio for the epidemic in Vietnam. We hope that the data presented here contribute to the epidemiology of COVID-19 worldwide.

## Methods

### Case identification

The data used in the present analysis were collated from two publicly available datasets: the Wikipedia [4] and the Johns Hopkins Dashboard system [3, 5]. The Wikipedia dataset includes data pertinent to date of diagnosis, gender, age, hospital admission, treatment status for each individual. In some cases, the source of infection was also ascertained. The dataset itself were collected from the information provided by the Vietnam Ministry of Health. The Johns Hopkins’ Dashboard includes the incidence of COVID-19 infection in each day for all countries in the world. This work was conducted using public datasets, and hence was determined not to be human subjects. All data were constructed in an anonymous fashion.

### Data analysis

An epidemiologic curve was constructed from the daily number of confirmed cases using a standard method [6]. The incidence of COVID-19 cases was analyzed by age group and gender. In addition, the reproduction ratio (*R*0) was estimated by the method described by Cori et al [7] using a previously published time interval [8]. The 95% confidence interval for *R*0 was determined by bootstrap method, with 10,000 bootstrap samples. The global force of infection (i.e., the per-capita rate at which susceptible individuals get infected) was also determined from observed data. All analyses were conducted using the R Statistical Environment [9].

## Results

Between 23/1/2020 and 10/4/2020 (79 days), 257 individuals have been confirmed to have been inflected with SARS-cov-2. The epidemiologic curve (**Figure 1**) shows two waves of outbreaks: the first wave had occurred between 23/1/2020 and mid-February 2020 with sporadic cases; and the second, larger wave of infections was observed from the end of February 2020.

**Figure 1:**
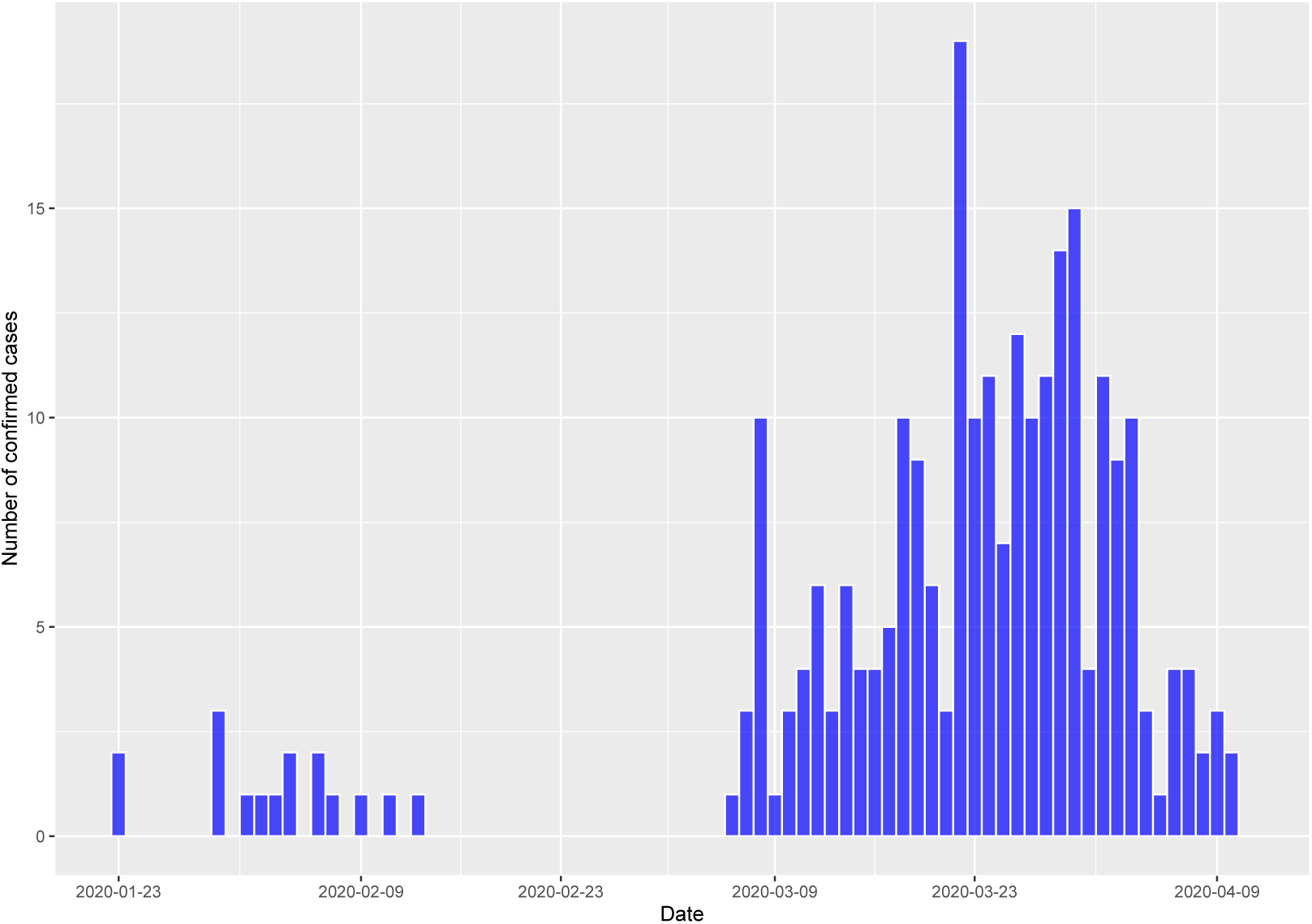
Number of cases who have been confirmed to be infeced by SARS-Cov-2 in Vietnam from 23/1/2020 to 10/4/2020.

The estimated reproduction ratio for the epidemic was 1.08, with 95% confidence interval being between 0.95 and 1.24 (**Figure 2**). The time-variant force of infection shows that the percapita rate at which susceptible individuals acquire infection has been in a downward trend since early April (**Figure 3**).

**Figure 2:**
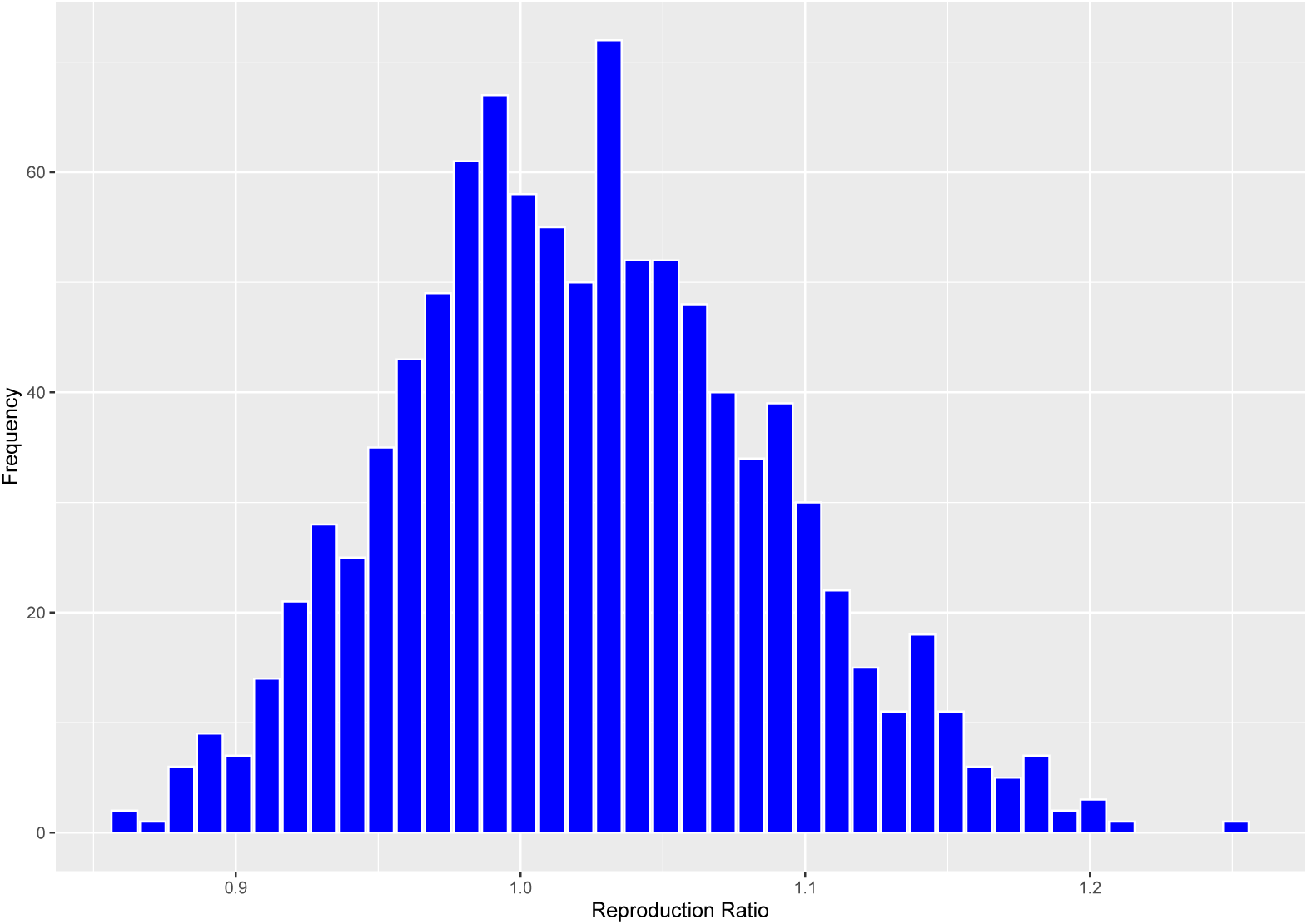
Distribution of the reproduction ratios (R_0_) based on 10,000 bootstrap samples. The median of R_0_ was 1.02 (95% confidence interval: 0.95 to 1.24)

**Figure 3:**
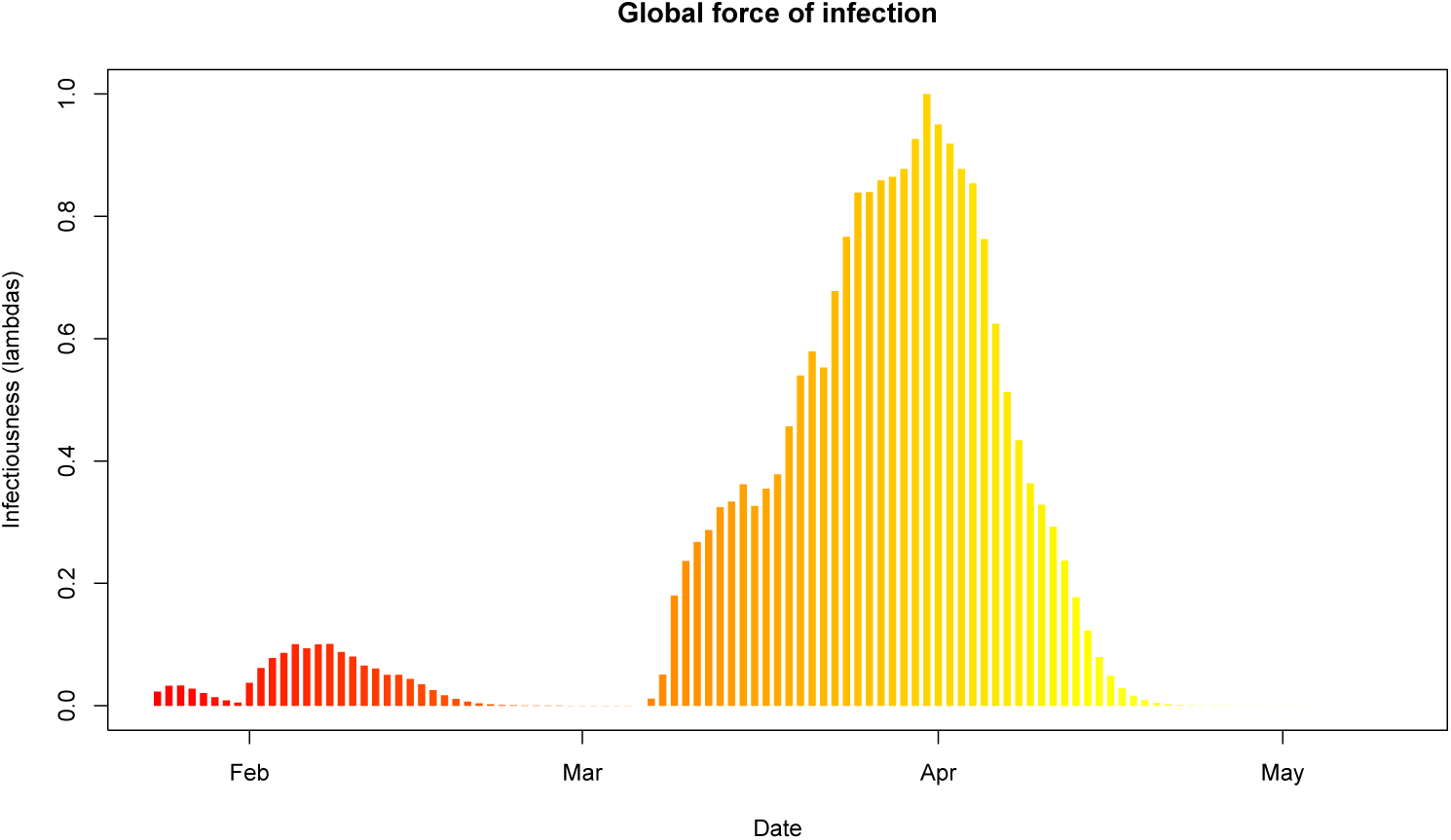
Force of infection for COVID-19 epidemic in Vietnam.

Just over half (54%; n = 139) of the confirmed cases were women. The median age was 30, with range being 3 months of age to 88 years. Almost 78% (n = 196) of patients aged 49 years or younger, and only 10% (n = 26) patients aged 60 and older (**Figure 4**).

**Figure 4:**
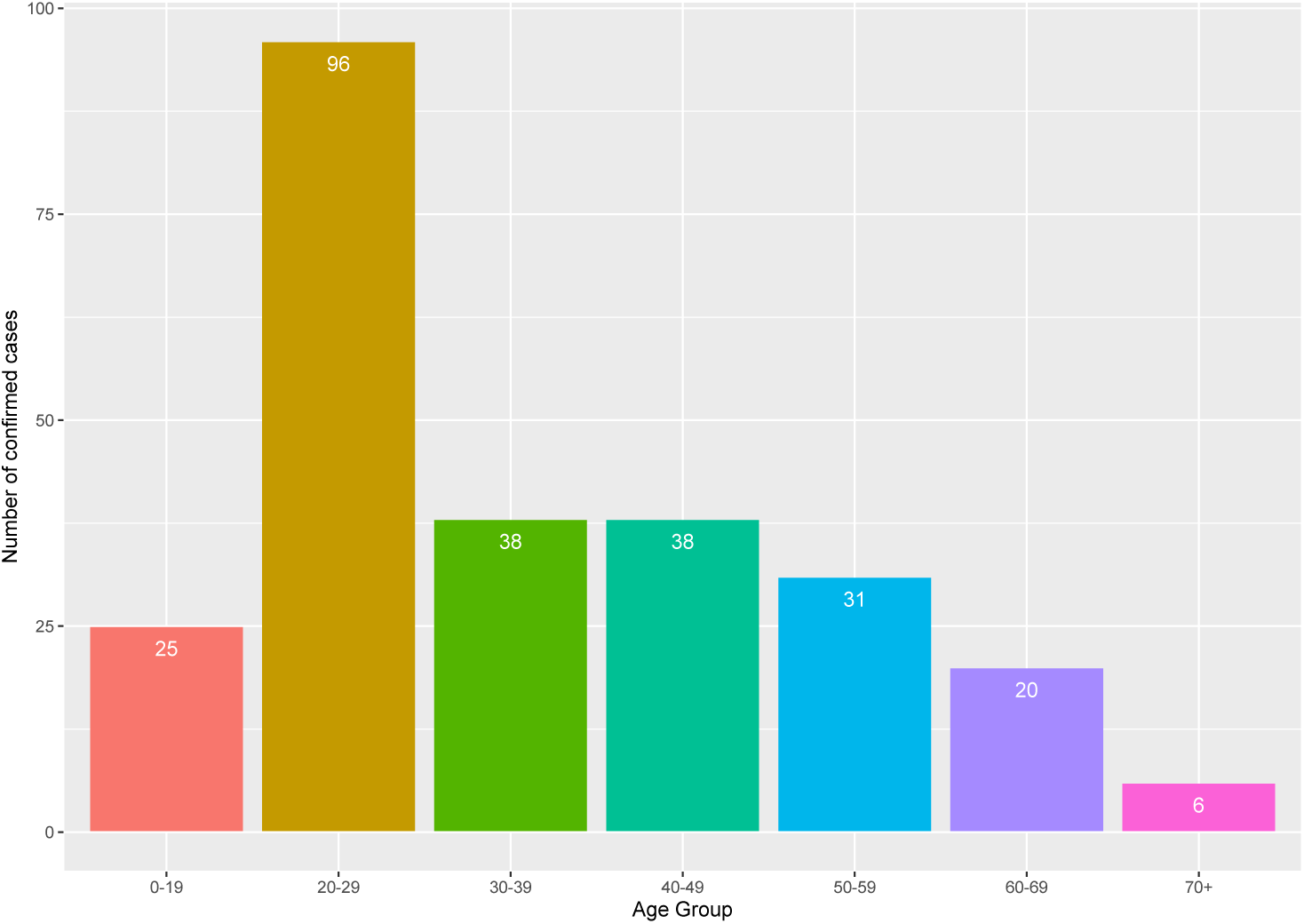
Distribution of age among 254 COVID-19 patients who age was ascertained.

Among the 257 patients, approximately 20% (n = 52) were international tourists or overseas workers. Most overseas patients were from the United Kingdom (n = 19), France (n = 5), the United States (n = 5). Among the 205 Vietnamese patients, 58% (n = 119) were students and/or workers returning from overseas. Nine patients reported that they had visited China prior to the confirmed infection (**Table 1**).

**Table 1:** Characteristics of 257 patients infected with SARS-Cov-2 in Vietnam.

| **Characteristics** | **Number of cases (%)** |
| --- | --- |
| **Gender** |  |
| Women | 136 |
| Men | 119 |
| **Age group** |  |
| 0 - 19 | 25 (9.8) |
| 20 - 29 | 96 (37.8) |
| 30 - 39 | 38 (15.0) |
| 40 - 49 | 38 (15.0) |
| 50 - 59 | 31 (12.2) |
| 60 - 69 | 20 (7.9) |
| 70+ | 6 (2.4) |
| **Returning from overseas** |  |
| Yes | 155 (60.3) |
| No | 139 (39.7) |
| **Nationality** |  |
| Vietnamese | 205 (80.7) |
| UK | 19 (7.5) |
| USA | 5 (2.0) |
| France | 5 (2.0) |
| Others | 23 (7.8) |
| **Treatment status** |  |
| Being treated | 110 (43.3) |
| Discharged | 144 (56.7) |

Note: Data were collated between 23/1/2020 and 10/4/2020.

Until 10/4/2020, approximately 57% (n = 144) patients have been recovered and discharged from hospitals. The rest (43%) are still being treated in various hospitals in the country. There was no statistically significant (*P* = 0.34) difference in age between those recovered (35 years) and those being treated (37 years). There have been no deaths reported since the outbreak.

## Discussion

The COVID-19 pandemic has affected millions of people globally. A key feature of the pandemic is that the incidence of COVID-19 infections varied remarkably between countries and territories, with cases per million people being high in Europe (e.g., Switzerland, Spain, Italy), but low in Asia (e.g., China, Singapore, Taiwan, Thailand). An analysis into the difference between countries may provide insights into the understanding of the pandemic. In this analysis, we have shown that the incidence of COVID-19 infections in Vietnam also low (around 2.7 per 1 million people) compared with that in European populations. This finding deserves some elaboration.

The epidemiologic profile of COVID-19 patients in Vietnam is very different from China and European countries. In China, male patients accounted for 51% of total patients, but in the present study, men were the minority group, accounting for 46% of total [10]. Moreover, the patients in this study were younger than those in China. Indeed, the median age of all patients was 30 years, which is younger that those in China (∼51 years), Germany (49), France (62.5), and Italy (62) [11]. In this study, only 10% of patients aged 60 years and older, and this figure represents one-third the proportion observed in China [12].

A majority of patients (67%) in this study were either international tourists or Vietnamese returning from overseas. While overseas visitors were typically in their 40s, most of the returnees were of young age, typically 20-30 years old, and that explains the young age in this study. The present dataset did not record clinical characteristics of patients, and as a result, it was not possible to ascertain the prevalence of comorbidities in the patients.

A positive feature of this sample of patients was that almost 60% of them have been recovered, and this proportion was not related to either age or gender. Based on the mortality rate in Wuhan [10], the expected number of deaths for this sample is between 2 to 5, but in reality there have been no deaths recorded among the patients, and this is a remarkable outcome.

A key parameter in any epidemic is the reproduction ratio. The estimated reproduction ratio in this study was between 0.95 and 1.24, which is relatively low compared with the estimate in China which peaked at 3.08 [13]. Moreover, a positive sign is that the force of infection has declined in recent weeks, suggesting that the COVID-19 epidemic in Vietnam is under controlled.

The present results should be interpreted in the context of limitations. It can be argued that the reliance on wikipedia data is a weakness, because we could not directly ascertain the infection. Because there were no clinical data, we could not evaluate the severity and treatment outcome of patients. It is important to note that the epidemic is still going on, and data are continuously evolved by the epidemic course; thus all estimates should be treated as temporary and time-dependent.

In Vietnam, the government has been proactive since the first case identified. A series of public health measures have been introduced to control the epidemic. These measures included stringent exit screening at airports, limited travel ban, application of information technology in the identification of potential cases, social distancing, and limited lockdown in major cities. These measures have been supported and folowed by the public at large.

In summary, this analysis has shown that COVID-19 patients in Vietnam were largely imported cases in overseas tourists and young students and workers who had returned from overseas. The low estimated reproduction ratio and force of infection show that the epidemic is on the downward trend, suggesting that the government’s strategy of containment has been effective.

## Data Availability

All data are available on reasonable request.

## Statement of Authorship

- Conception or design of the work: TVN
- Data collection: HGN and TVN
- Data analysis: HGN
- Drafting the article: TVN
- Final approval of the paper: HGN, TVN

